# Factors associated with adverse outcomes among patients hospitalized at a COVID-19 treatment center run by Médecins sans Frontières in Herat, Afghanistan

**DOI:** 10.1101/2023.02.15.23285976

**Authors:** Ana Klein, Mathieu Bastard, Hamayoun Hemat, Saschveen Singh, Bruno Muniz, Guyguy Manangama, Amber Alayyan, Abdul Hakim Tamanna, Bashir Barakzaie, Nargis Popal, Mohammad Azeem Zmarial Kakar, Elisabeth Poulet, Flavio Finger

## Abstract

**Background:** Though many studies on COVID have been published to date, data on COVID-19 epidemiology, symptoms, risk factors and severity in low- and middle-income countries (LMICS), such as Afghanistan are sparse.

**Objective:** To describe clinical characteristics, severity, and outcomes of patients hospitalized in the MSF COVID-19 treatment center (CTC) in Herat, Afghanistan and to assess risk factors associated with severe outcomes.

**Methods:** 1113 patients were included in this observational study between June 2020 and April 2022. Descriptive analysis was performed on clinical characteristics, complications, and outcomes of patients. Univariate description by Cox regression to identify risk factors for an adverse outcome was performed. Adverse outcome was defined as death or transfer to a level 3 intensive care located at another health facility. Finally, factors identified were included in a multivariate Cox survival analysis.

**Results:** A total of 165 patients (14.8%) suffered from a severe disease course, with a median time of 6 days (interquartile range: 2-11 days) from admission to adverse outcome. In our multivariate model, we identified male gender, age over 50, high O2 flow administered during admission, lymphopenia, anemia and O2 saturation <=93% during the first three days of admission as predictors for a severe disease course (p<0.05).

**Conclusion:** Our analysis concluded in a relatively low rate of adverse outcomes of 14.8%. This is possibly related to the fact, that the resources at an MSF-led facility are higher, in terms of human resources as well as supply of drugs and biomedical equipment, including oxygen therapy devices, compared to local hospitals. Predictors for severe disease outcomes were found to be comparable to other settings.

## Introduction

### Covid-19

Since the initial outbreak of the coronavirus disease (COVID-19) in December 2019 in Wuhan, China, SARS-CoV-2 has spread across the globe affecting millions of people. As of July 2022, the number of past and current infections reported to the WHO rose to more than 550 million cases with more than six million cumulative deaths.^1^

Though most cases are mild and do not require hospitalization, approximately 14% of patients recorded in the literature experience a severe and 5% a critical course^2, 3^. This, however, highly depends on the age structure and prevalence of underlying risk factors within the population. In populations with a younger age structure, such as Afghanistan, the proportion of recorded severe and critical disease cases tend to be lower.^4^ Common complications described in studies of patients with a severe disease course include acute respiratory distress syndrome (ARDS) or cardio- and cerebrovascular complications, due to the pro-coagulant nature of the disease.^5-9^

After admission to hospital, published mortality rates range widely between 2% and 60%, depending on various factors such as the type of care facility (e.g., equipped for critical care or not), hospital equipment and number and qualification of staff.^10-15^ Multiple meta-analyses have estimated the in-hospital mortality to be around 17% (95% CIs ranging from 12.7% to 22.7%)^16, 17^

Many studies have researched predictors for in-hospital mortality. Identified factors have included: higher age, male gender, low oxygen saturation at admission, tachypnea and various laboratory determinants such as lymphopenia, low hemoglobin levels, elevated c-reactive protein (CRP), lactate dehydrogenase (LDH) and urea, hyponatremia, hyperkalemia and abnormal coagulation parameters.^11, 18-25^

### Rationale

From China the virus spread first to high-income countries, followed shortly by low- and middle-income countries (LMIC). Due to the weaker socioeconomic status, fragile health-systems and infrastructure of these countries, the impact of COVID-19 on the population and health systems raised concerns. The situation in LMICs may lead to a different profile of disease severity and finally potentially higher risk of death in individuals with risk factors for severe disease, due to a lack of medical infrastructure, skilled staff, intensive care capacity and biomedical equipment or otherwise well-functioning services that become quickly overwhelmed.^14^ Furthermore, while LMICs tend to have a younger age structure,^26, 27^ which has been identified as a possible protective factor,^28^ non-communicable diseases such as diabetes and chronic vascular disease are on the rise in many LMICs and are often poorly controlled, or undiagnosed and consecutively left untreated, predisposing to higher risk of complications.^29-32^

From both a public health and a clinical perspective, it is therefore vital to determine key predictors for unfavorable disease outcomes, in order to better estimate the likely burden on the health system and the resources required for future waves of COVID-19, to aid physicians in triaging patients appropriately and allocating valuable resources such as oxygen therapy to those in greatest need.

The first case of COVID in Afghanistan was detected in February 2020 in Herat.^33^ Nationally, as of July 2022, approximately 180,000 confirmed cases and 7700 deaths from COVID have been reported to the WHO. It is, however, estimated that the actual numbers are much higher due to persistent limitations in capacity of laboratory and surveillance infrastructure.^1, 34^ The first wave of disease is hypothesized as being linked to the large influx of Afghan refugees returning from neighboring Iran, which was heavily affected in the early stages of the pandemic.^35, 36^ As of July 2022, Afghanistan has been hit by four waves, the first spanning from April to June 2020, the second from October 2020 to December 2020, the third from April 2021 to August 2021 and the fourth and most recent wave from January to April 2022.^37^

Since the onset of the pandemic, Médecins Sans Frontières (MSF) has been working in many affected countries, with interventions ranging from basic community education and health worker trainings, to setting up mobile clinics, and COVID-19 treatment centers (CTC) of varying capacity and levels of care. In Herat, Afghanistan, MSF set up a CTC which opened in July 2020. Here, we report on the disease characteristics, severity, and outcomes among COVID-19 patients admitted to the CTC.

## Methods

### Study design

The study follows a mixed prospective and retrospective observational design. Data collection was performed between 26^th^ of June 2020 and 14^th^ of March 2022. The prospective component began after approval of the protocol by the institutional Review Board of Afghanistan on the 16^th^ of August 2020. All data from before this time was collected retrospectively from clinical patient files.

### Study site

Study site is the MSF CTC in Herat, Afghanistan, a major city (estimated population of 600’000)^38^ and the regional capital of the Western Region and provincial capital of the Herat province (estimated population of 2.2M)^38^. The CTC was initially providing basic level 1 ICU care with the provision of standard oxygen therapy, later upgrading to level 2 ICU capacity from December 2020 with the arrival of non-invasive ventilation equipment. The CTC admitted patients corresponding to the MSF definition (and WHO’s initial 2020 definition) of moderate and severe COVID-19 disease, as identified at the MSF-run COVID-19 triage located at the Herat Regional Hospital, which acts as the corner stone of the COVID-19 care system in Herat. The CTC initially opened in June 2020. It was temporarily closed after first and second waves of COVID-19, to reopen at the start of the next wave. It did however remain open between wave three and four due to continuous presentation of cases meeting admission criteria, until its definitive closure in April 2022. It is important to note however that the MSF triage remained open both in between and throughout successive waves, this was part of MSF’s surveillance strategy and to ensure that patients presenting with COVID-19 between major waves could be identified and either isolated as outpatients or referred for inpatient isolation and management. The MSF CTC did not initially admit patients presenting in a critical state since there was no possibility of high flow non-invasive ventilation or intubation. Instead, critical patients were sent to Shaidayee hospital, a CTC run by a local NGO on behalf of the MoH. From Dec 2020 with the arrival of High Flow Nasal Oxygen therapy (HFNO), MSF teams kept patients meeting MSF criteria for critical disease (i.e. those requiring high flow non-invasive ventilation could be managed if deemed appropriate by MSF clinicians). Other treatments included antibiotics, antipyretics, steroids, and anticoagulants and treatments for any co-morbidities according to MSF protocol.

PCR (polymerase chain reaction) testing was not done for all patients due to lack of laboratory capacity in Herat. The national policy, which recommends testing of suspected cases over 50, severe cases (i.e., those requiring oxygen and hence admission), health care workers and pregnant women with symptoms, was applied to prioritize testing. Antigenic Rapid Diagnostic Tests (RDTs) were used to complement when available.

### Study population

The study population consists of all clinically suspected or lab-confirmed COVID-19 patients admitted to the CTC who have consented to their data being used or met the exemption criteria (see Ethical considerations).

Inclusion criteria were designed as: Clinically suspected or lab-confirmed COVID-19, and consent to participate in the study, and with outcome either discharge to home, transfer to ICU or death.

### Data collection

Routinely collected data included the patients’ medical history, clinical examination including vital signs at admission, lab results, including COVID-19 RT-PCR done at the Herat Regional Reference Laboratory, antigen RDTs, baseline blood test results and other clinically indicated tests upon the physician’s discretion where available (e.g., serological testing for human immunodeficiency virus (HIV), X-ray, or pregnancy tests). Patients were continuously monitored, and their vital signs documented throughout the day. In general, two daily values of vital signs were entered into the database (approximately at 8am and 6pm). Data on treatments, outcomes and complications were also collected. More information can be found in the supplementary table.

Study data were collected from patient files as documented by physicians and nurses during admission, stay and discharge. Study data were collected and managed using REDCap electronic data capture tools hosted at Epicentre, Paris.^39, 40^. All identifying information was removed, so that only deidentified data with a patient identification number is used for the statistical analysis.

Data was collected throughout four waves of the pandemic, whenever the CTC was admitting patients:

1. First Wave: admissions from the 26^th^ of June to the 20^th^ of September 2020,
2. Second Wave: admissions from the 1^st^ of December 2020 to the 1^st^ of March 2021
3. Third Wave: admissions from the 8^th^ of June 2021 to the 25^th^ of October 2021
4. Fourth Wave: admissions from the 26th of October 2021 to the 14^th^ of March 2022

### Descriptive analysis

Continuous variables were described by median and interquartile range (IQR), categorical data by counts, proportions and 95% confidence intervals (95% CI). Descriptive analysis was performed on sociodemographic data, clinical characteristics, complications, and outcomes.

### Severity at admission

Severity was assessed by the physicians in charge upon admission and based on the MSF COVID-19 guidelines (Table 1).

**Table 1:**
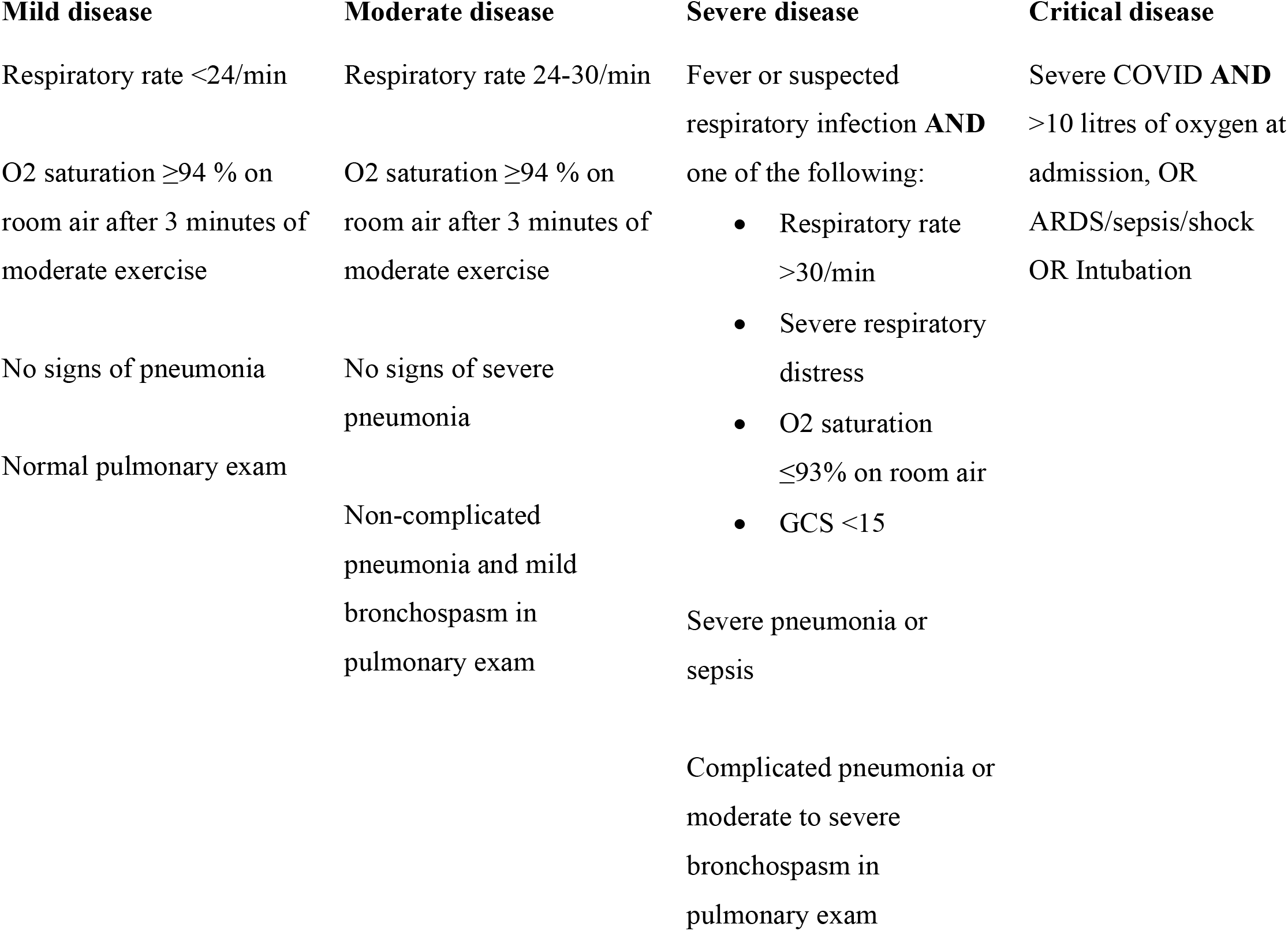
Disease severity at admission based on the MSF COVID guidelines

| Mild disease | Moderate disease | Severe disease | Critical disease |
| --- | --- | --- | --- |
| Respiratory rate <24/min | Respiratory rate 24-30/min | Fever or suspected respiratory infection **AND** one of the following: | Severe COVID **AND** >10 litres of oxygen at admission, OR ARDS/sepsis/shock OR Intubation |
| O2 saturation $\geq 94\%$ on room air after 3 minutes of moderate exercise | O2 saturation $\geq 94\%$ on room air after 3 minutes of moderate exercise | Respiratory rate >30/min<br>Severe respiratory distress<br>O2 saturation $\leq 93\%$ on room air<br>GCS <15 | |
| No signs of pneumonia | No signs of severe pneumonia |  |  |
| Normal pulmonary exam | Non-complicated pneumonia and mild bronchospasm in pulmonary exam | Severe pneumonia or sepsis |  |
|  |  | Complicated pneumonia or moderate to severe bronchospasm in pulmonary exam |  |

### Adverse outcomes

Since the CTC wasn’t equipped for level 3 ICU (intubation and mechanical ventilation), until dec 2020 patients in critical states were generally referred to Shaidayee hospital, which had a level 3 ICU and was run by a local NGO on behalf of the MoH. From Dec 2020 with arrival of HFNO, MSF teams had capacity to admit patients requiring higher flow of O2 and thus reducing the need to refer. The outcomes of patients referred to Shaidayee could unfortunately not be determined on an individual basis, but anecdotal evidence states that the mortality among the patients referred was high.

Thus, for this study, adverse outcome was defined as death or transfer to the level 3 ICU at Shaidayee hospital, while a mild disease course was defined as discharge to home or referral to a convalescence unit.

### Cox regression

In a second step uni- and multivariable Cox regression with time dependent co-variates was performed to identify risk factors for adverse outcome, with death or transfer to a level 3 ICU vs discharge to home as the dependent variable. Independent covariates included basic patient demographics, vaccination status, COVID-19 test result, clinical information at admission, laboratory parameters and vital signs. Vital signs were included as time dependent co-variates. To adequately address our main research question, we only included data from the first three days after admission. Time at risk was calculated as the time between admission and outcome. To increase clinical relevance, we decided to categorize our variables and set clinically relevant cutoffs, which were as follows: Age > 50 years, hemoglobin <12 g/dl, lymphocytes <500 10^3^/μl and O2 saturation ≤93%.

As lymphocytes were collected as % of WBC, we transformed this variable into absolute values by multiplying with the median number of WBC over all measurements. Due to discrepancies within the database and a too high proportion of missing values, underlying comorbidities were not included in the analysis.

After performing the univariate analysis, we selected variables based on their statistical significance (p-value <0.1), their clinical relevance and completeness of data in descending order of importance to be included in our multivariate model. Adjusted hazard ratios were expressed with 95% confidence intervals (CI) and an alpha level of 5%. All analyses were performed using R 4.1.2 (The R Foundation for Statistical Computing, Vienna, Austria).

### Ethical considerations

The research described here has been conducted according to the principles expressed in the Declaration of Helsinki. This study was approved by the Institutional Review Board of the Afghan National Public Health Institute (Protocol A.0820.0214, 12 August 2020) and by the MSF Ethical Review board (Protocol 2043a, 20 August 2020). All patients included in the study either verbally consented for their deidentified data to be used or met the exemption criteria approved by the Ethical Review Boards (patient discharged before the study started and thus included retrospectively OR patient deceased before the verbal consent could be taken). Verbal consent was warranted due to widespread illiteracy and since the study presents minimal risk to participants and does not include any procedure for which written consent is normally required.

## Results

### Inclusions

During the entire period of operation, a total number of 1428 patients were admitted to the CTC. After exclusion of 315 patients who did not meet the inclusion criteria, a total of 1113 clinically suspected or serologically confirmed COVID-19 patients were included in the study. Of these, 99 patients were included in the retrospective part (discharge before the study start on 16.08.2020) and thus exempt from individual consent.

### Weekly admissions and outcomes

Figure 1 gives an overview of weekly admissions throughout the periods when the CTC was open, stratified by outcome. An average of just below 17 patients were admitted per week. During wave 1, 169 patients were included, during wave 2, 3 and 4, 278, 381 and 285 patients were admitted respectively.

**Figure 1:**
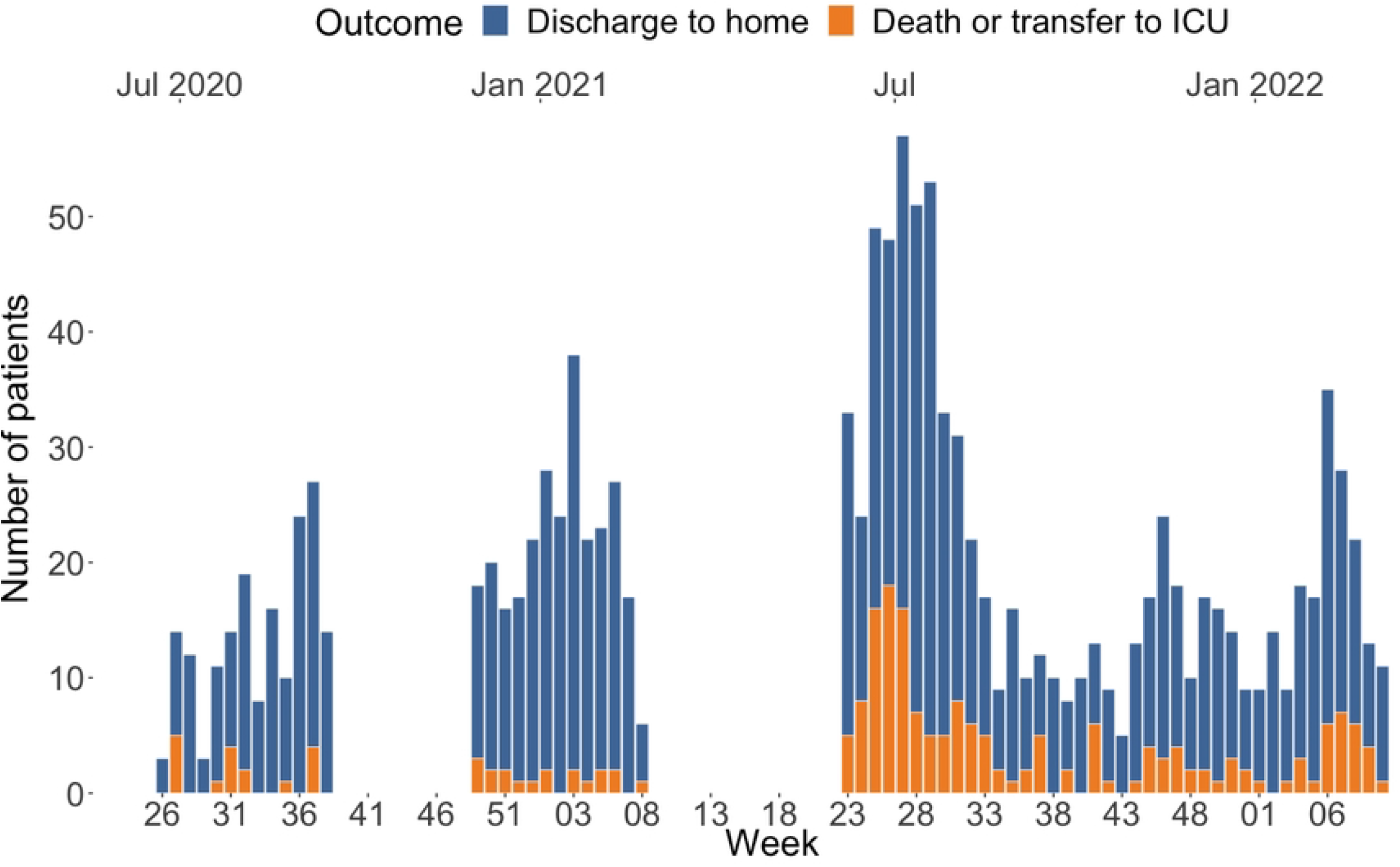
Weekly admissions stratified by outcome. Number of patients admitted to the CTC per week stratified by outcome. As can be seen in the graph, the CTC remained open between waved 3 and 4.

### Demographic and clinical characteristics

The median age of all patients was 60 years, with an IQR of 47 to 70 years and only slight variations between waves (Table 2). Of the 1109 patients for whom gender is known, 591 (53%) were female. Only during wave 3 the proportion of males was higher (59%).

**Table 2:**
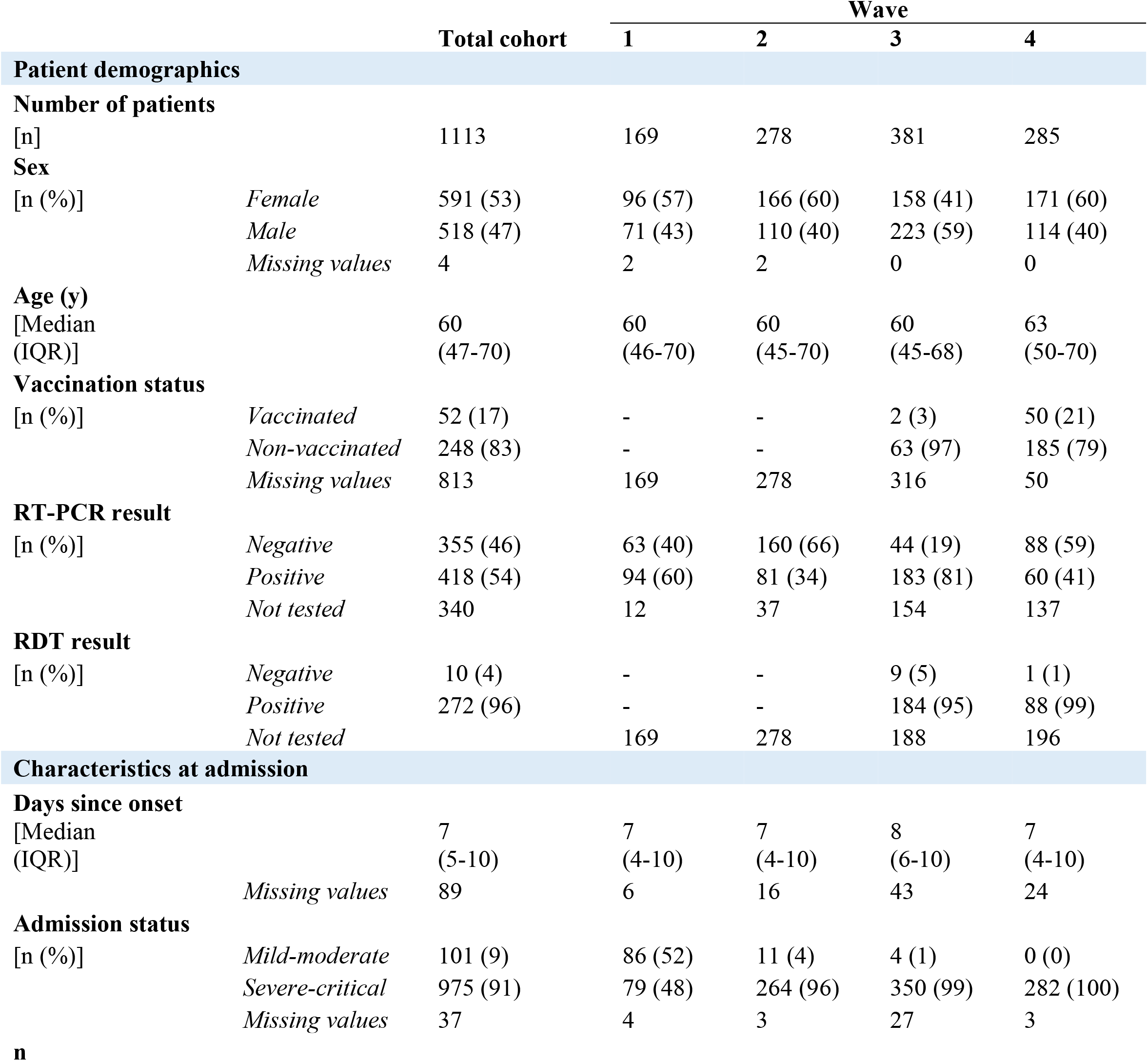
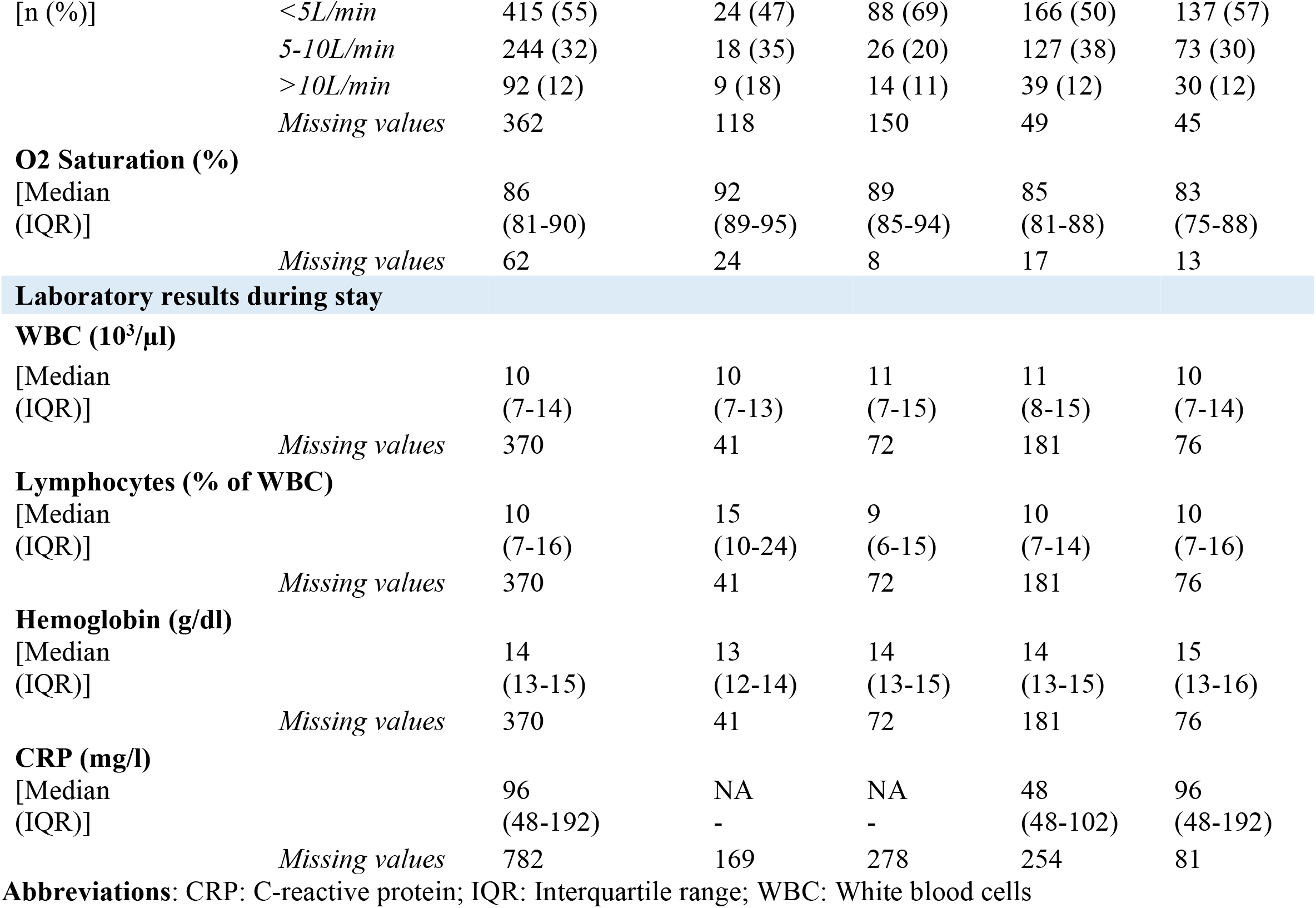
Sociodemographic and clinical characteristics

A total of 52 patients reported to be vaccinated against COVID-19, all of whom were hospitalized during the third and fourth waves (table 1, COVID-19 vaccination started in November 2021 in Herat). The proportion of vaccinated individuals was 17% when considering only patients admitted after vaccination first became available (52 out of 300 patients in total). The most common vaccine was Johnson & Johnson (40 patients, 77% of vaccinated), followed by AstraZeneca (3, 6%) and Sinopharm (1, 2%). The latter two vaccines require two doses, our patients however reported to have received one dose only. For 8 patients, data on the type of vaccine used was not available.

A total of 951 patients were tested for COVID-19, of these 109 patients were tested twice. The most commonly used test was RT-PCR, however in some cases antigenic RDTs were performed, which became available during the third wave. In total, 773 patients were tested with RT-PCRs and 282 with RDTs, while the choice of test was unknown for 5 cases. Of the patients tested with RT-PCR, 355 (46%) had a negative and 418 (54%) at least one positive RT-PCR or RDT result. Comparison of all four waves shows that the highest proportion of positive tests was during wave 3 with 183 patients (81%) testing positive at least once, compared to only 81 patients (34%) during wave 2 (Table 2). Almost all patients tested with RDTs had positive results, with only 10 out of 282 patients receiving a negative test result (4%).

Most patients were classified as severe or critical at admission (975 patients; 91% of total cohort), compared to only 101 patients (9%) who were mild to moderate. During the first wave 46% of patients were classified as severe upon admission, while this number increased continuously up to 100% during the fourth wave, which was partially due to limited bed capacity and stricter application of admission criteria.

All patients received oxygen at admission. 415 (55%) received under five, 244 (32%) five to ten, and 92 (12%) over ten liters of oxygen per minutes.

Median O2 saturation was 86% (IQR: 81-90%) at admission for the total cohort. Analysis stratified by wave showed that this number decreased through waves, with a median O2 saturation of 92% (IQR: 89-95%) at admission during the first and 83% (IQR: 75-88%) during the fourth wave (Table 2).

Basic blood laboratory analysis was performed in most patients. The most frequently performed analysis was a complete blood count with differential test, while other parameters such as CRP was tested only upon the physician’s discretion. If a patient received more than one blood analysis, results were averaged to facilitate analysis (Table 2).

During the first wave, the three most common symptoms documented at admission were fever, shortness of breath and cough. While cough and shortness of breath remained among the top three reported symptoms for the following two waves, presentation with fever became continuously less frequent. The third most frequently reported symptoms for waves 2 and 3 were chest pain and headache respectively, while muscle and chest pain tied for the third position during the fourth wave. Abdominal and nasal symptoms were of less importance throughout all four waves (Figure 2).

**Figure 2:**
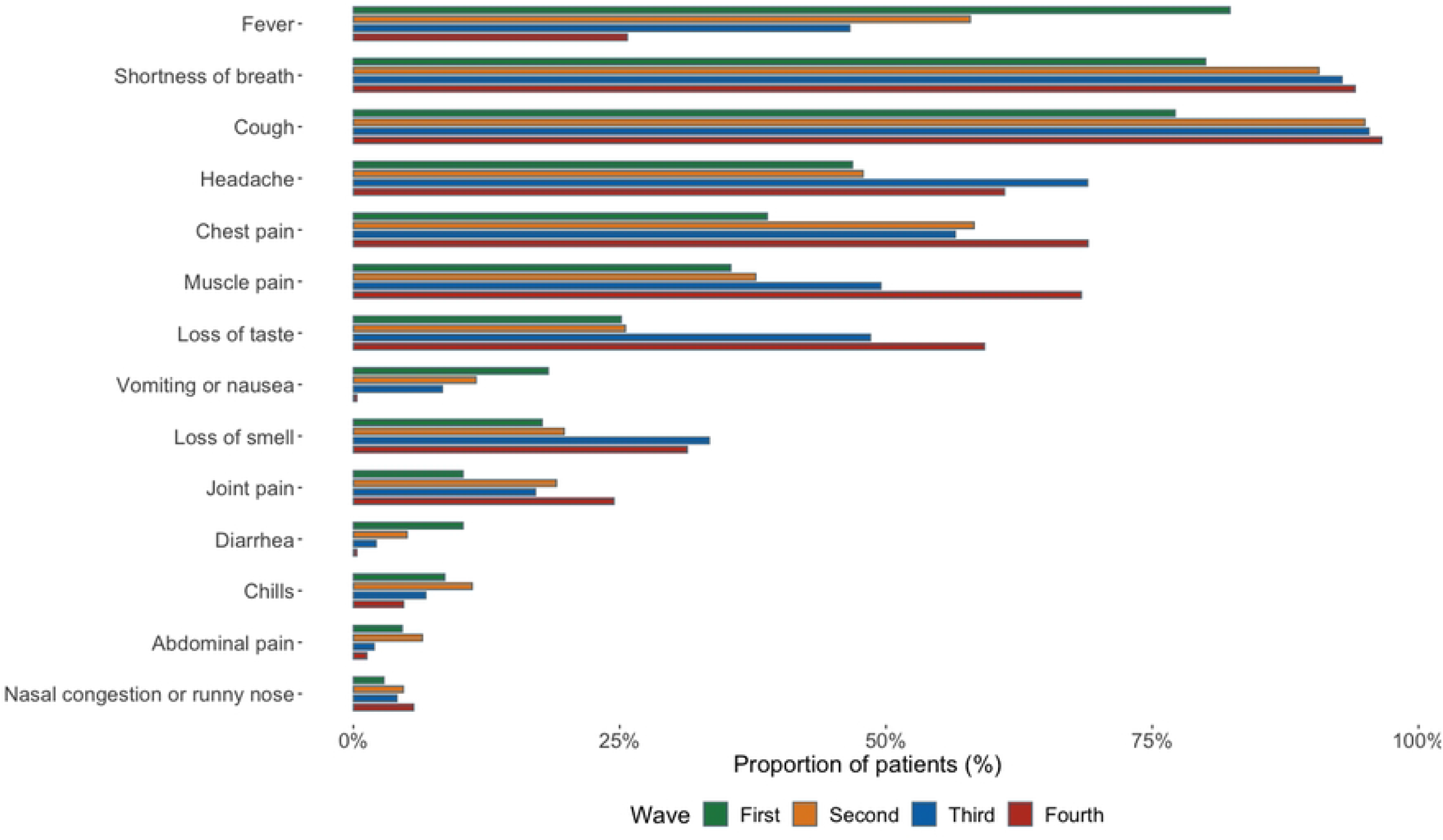
Most common symptoms of patients at admission stratified by wave

### Vital signs

We analyzed the evolution of O_2_ saturation and systolic and diastolic blood pressure over the first ten days after admission stratified by outcome (Figure 3), including a linear trend line. Overall, the median O_2_ saturation levels of patients with a more severe outcome were lower than in those with a positive outcome. While the trend line of both groups decreases over time, the decrease is steeper in patients with an adverse outcome. Median O_2_ saturation levels at day 2 were 94% (IQR: 92-96%) in patients with a positive outcome vs 91% (IQR: 88-94%) in patients with an adverse outcome. On days 5 and 10 saturation levels were 94 vs 89 % (IQR: 91-95% and 85-93%) and 93 vs 88% (90-94% and 84-91%) respectively.

**Figure 3:**
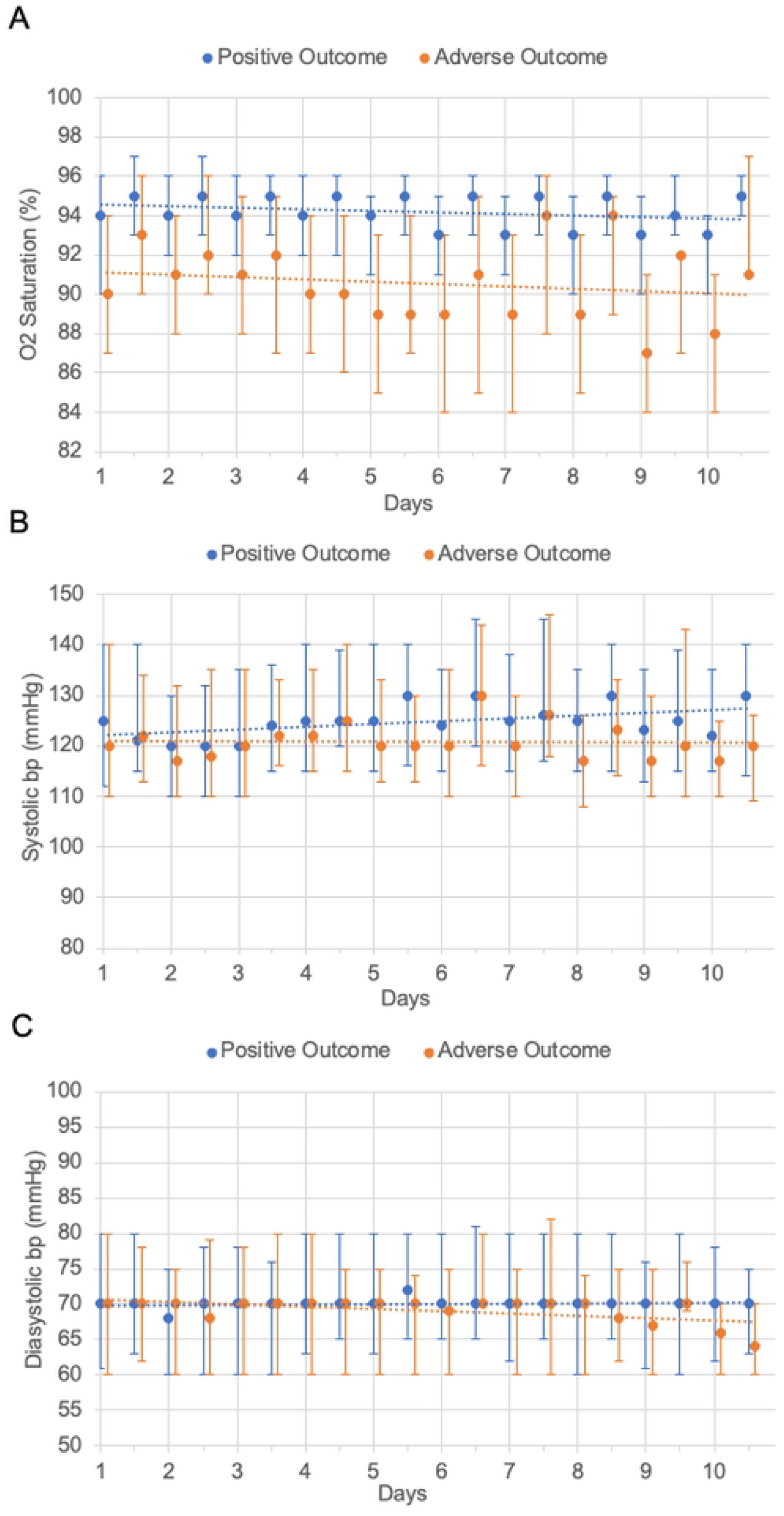
Evolution of patients’ vital signs over the first ten days after admission A: O2 Saturation over time; B: Systolic blood pressure over time; C: Diastolic blood pressure over time All graphs include median values, IQR and a linear trendline. Values for patients with a positive outcome are shown in blue, orange stands for patients with an adverse outcome. **Abbreviations**: bp: blood pressure

For the evolution of systolic and diastolic blood pressure, the difference according to outcome is less pronounced (Figure 3B and C). Regarding systolic blood pressure, patients with an adverse outcome have slightly lower blood pressure values than those with a positive outcome. While the trend line of the first group remains stable, the trend line of the latter shows a slight increase of blood pressure values over time. Regarding diastolic blood pressure, values are at an equal level in both groups. Note the important number of values far over the normal range. Also note that this descriptive analysis of vital signs suffers from right-censoring since patients who were discharged do not contribute to the averages of subsequent days.

### Complications and outcomes

A total of 78 patients (7.0 %) experienced one or more complications during their hospital stay, with the largest proportion of patients experiencing complications during the first wave (18 patients; 10.7%), and the smallest during the second (8 patients; 2.9%, table 3). The three most frequent were pneumological complications such as respiratory failure, ARDS, and pneumonia (37, 21 and 19 patients or 3.3, 1.9 and 1.7% of all patients respectively), followed by cardiac problems such as heart failure or shock.

**Table 3:** Complications and outcome

| Characteristic |  | Total cohort | Wave |  |  |  |
| --- | --- | --- | --- | --- | --- | --- |
|  |  |  | 1 | 2 | 3 | 4 |
| Complications |  |  |  |  |  |  |
| Respiratory failure |  |  |  |  |  |  |
| [n (%)] |  | 37 (3.3) | 5 (3.0) | 5 (1.8) | 18 (4.7) | 9 (3.2) |
| ARDS |  |  |  |  |  |  |
| [n (%)] |  | 21 (1.9) | 1 (0.6) | 0 (0.0) | 14 (3.7) | 6 (2.1) |
| Pneumonia |  |  |  |  |  |  |
| [n (%)] |  | 19 (1.7) | 9 (5.3) | 3 (1.1) | 5 (1.3) | 2 (0.7) |
| Heart failure |  |  |  |  |  |  |
| [n (%)] |  | 13 (1.3) | 3 (1.8) | 2 (0.7) | 6 (1.6) | 2 (0.7) |
| Shock |  |  |  |  |  |  |
| [n (%)] |  | 10 (0.9) | 1 (0.6) | 0 (0.0) | 5 (1.3) | 4 (1.4) |
| Other complications* |  |  |  |  |  |  |
| [n (%)] |  | 21 (1.9) | 3 (1.8) | 2 (0.7) | 5 (1.3) | 11 (3.9) |
| Any complications** |  |  |  |  |  |  |
| [n (%)] |  | 78 (7.0) | 18 (10.7) | 8 (2.9) | 31 (8.1) | 21 (7.4) |
| Outcome |  |  |  |  |  |  |
| [n (%)] | ICU/Death | 165 (14.8) | 15 (8.9) | 19 (6.8) | 89 (23.4) | 42 (14.7) |
|  | Discharge | 948 (85.2) | 154 (91.1) | 259 (93.2) | 292 (66.6) | 243 (85.3) |
| Days to outcome |  |  |  |  |  |  |
| [Median (IQR)] | ICU/Death | 6 (2.0-11.0) | 6.1 (0.1-9.1) | 3.0 (1.5-7.5) | 6.1 (3.1-10.1) | 3.5 (2.0-10.0) |
|  | Discharge | 5 (2.1-6.1) | 3.1 (2.1-4.1) | 3.0 (2.0-6.0) | 5.1 (3.1-8.1) | 4.0 (3.0-7.0) |

\* All complications occurring in less than 10 patients \*\* Patients may have had more than 1 complication
**Abbreviations:** ARDS: Acute respiratory distress syndrome; IQR: Interquartile range

Within the complete cohort a total of 165 patients (15%) experienced an adverse outcome, while 948 patients (85%) were discharged to home. The lowest proportion of adverse outcomes occurred in wave 2 (19 patients; 6.8%), while the highest was documented during wave 3 (89 patients; 23%, table 3). Median time to discharge to home was 5 days (IQR: 2.1-6.1 days) for the total cohort with variations between 3 days (IQR: 2.0-6.0 days) in wave 2 and 5 days (IQR: 3.1-8.1 days) in wave 3. Median time to adverse outcome was generally one day longer (Table 3).

### Multivariable survival analysis

To identify possible factors associated with a severe disease course, we performed uni- and multivariate survival analysis using Cox proportional hazard models.

In our univariate model male gender, a severe to critical status at admission, higher O2 flow at admission, an increase in white blood cells, anemia, O2 saturation < 94% and higher systolic and diastolic blood pressure were significantly associated with an increased risk of adverse outcome (p<0.05).

Our multivariate model included gender, age, wave, O2 flow at admission, lymphocytes, hemoglobin and O2 saturation over the first three days. Apart from epidemic wave and lymphopenia, all variables showed a significant association with the outcome (p<0.05).

Male gender and age over 50 both were associated with an approximately twofold increase in risk of an adverse outcome with a HR of 1.92 (95% CI: 1.14-3.24; p-value: 0.015) and 2.28 (95% CI: 1.14-4.56; p-value: 0.020) respectively. A higher O_2_ flow at admission was also associated with an increase in risk of an adverse outcome, with a HR of 2.52 and 5.19 for an oxygen flow between 5 and 10 liters and over 10 liters respectively (95% CIs: 1.32-4.79 and 2.62-10.29; p-values: 0.005 and <0.001 respectively). Furthermore, anemia with a hemoglobin level of less than 12 g/dl was associated with a statistically significant increase in risk (p<0.05) of an adverse outcome, while lymphopenia with <500 cells × 10^3^ /μl showed a trend towards an association with the outcome but was not significant with a p-value of 0.059 (Table 4). Regarding our time-dependent variable, an O_2_ saturation of under 94% within the first three days was associated with a twofold increase in risk of an adverse event, with an HR of 2.08 (95% CI: 1.14-3.81).

**Table 4:**
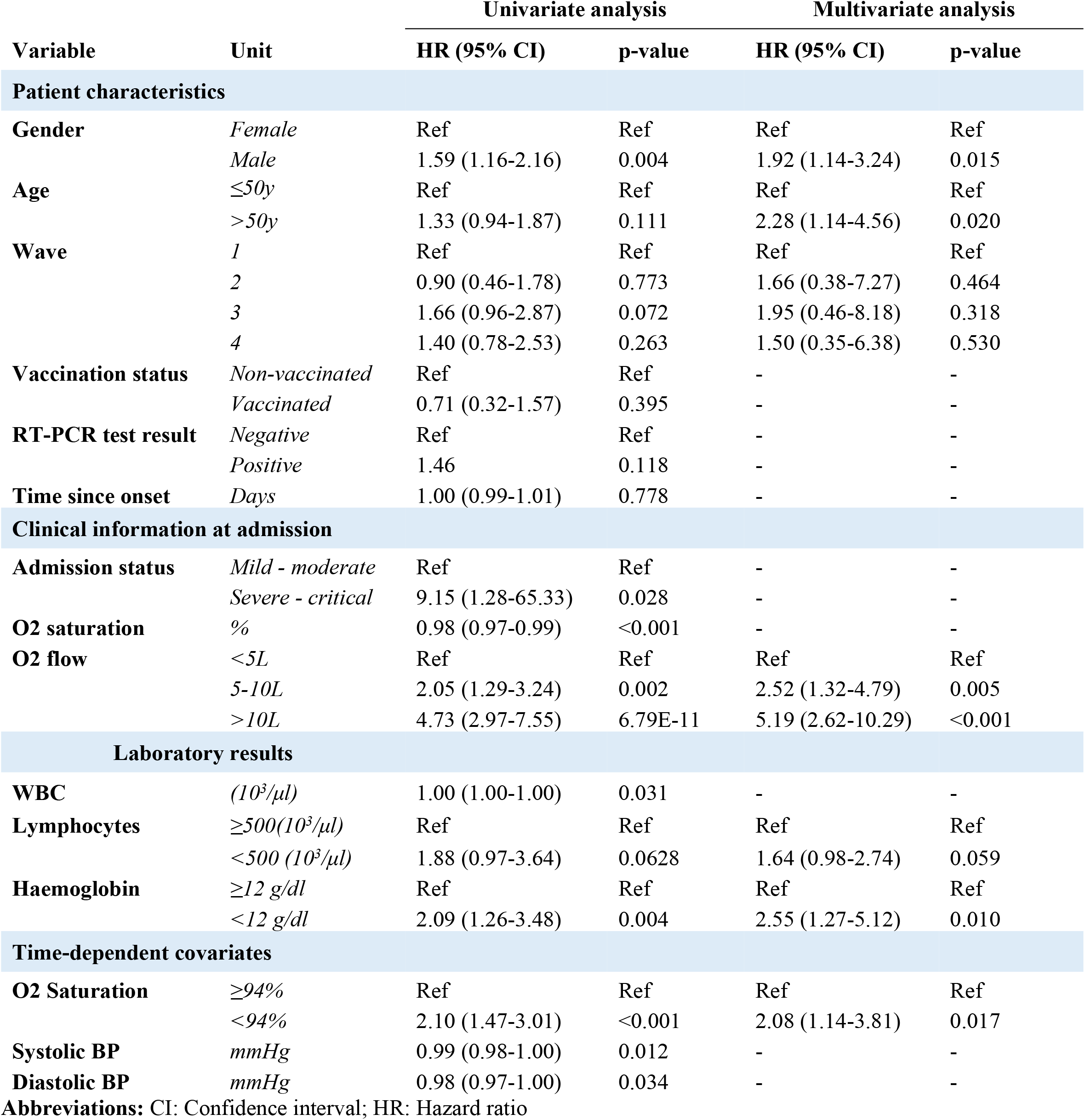
Univariate and multivariate Cox Proportional-Hazards Model with time-dependent covariates

## Discussion

Although COVID-19 is a novel disease that emerged only in late 2019, it has attracted a great amount of scientific attention, likely due to its rapid evolution into a global pandemic, leading to a cumulative death toll of over 6.3 million people worldwide(as of July 2022), not including additional mortality from other causes due to the secondary effects of the pandemic on health services.^1^ The evolution of the pandemic in LMIC was of particular concern due to the fragility and limited capacity of health systems and populations that are often subject to high prevalence of co-morbidities. However, the scientific literature on COVID-19 in LMIC in general remains sparse. For Afghanistan in particular, among other factors, this is most likely related to challenges in data collection and reporting, the persistent limitations in availability of testing and in access to health care services for a large part of the population. The recent political changes and subsequent retreat or downscale of programs of many humanitarian organizations probably also had an impact on the quality of the pandemic documentation. MSF has been present throughout the pandemic and maintained a triage service for suspect cases consistently even between waves and a multidisciplinary inpatient CTC service throughout each of the first four waves, this study offers a unique insight into the clinical presentation of COVID-19 in the context of Afghanistan.

Our analysis focuses on the description of the clinical characteristics, severity, and outcomes of hospitalized COVID-19 patients in Herat, Afghanistan. A total of 1113 patients were included in this study with a median age of 60 years (IQR: 47-70 years) and a slightly higher proportion of females, which is comparable with other cohorts throughout the globe, even though an inverse gender distribution is more common elsewhere.^13, 41-46^

The proportion of patients classified as severe or critical at admission increased from 46 to 100% over the course of the four waves. This is explained by the limited bed capacity and resulting variation in rigor in the application of the admission criteria.

The distribution of symptoms within our cohort was similar to that of other cohorts with the most common symptoms at presentation including cough, shortness of breath, fever, headache, and chest pain.^13, 42, 45-48^ Of note is the gradual change in the proportion of patients presenting with fever, which decreased from 82% during the first wave to 24% during the fourth wave, in line with other contexts.^49^ One large multicenter retrospective analysis on 21,461 unvaccinated Spanish COVID patients for example found that while fever was reported by 70-74% of patients during waves 1 and 2, this number decreased to 58.3% in successive waves.^50^ One possible explanation is the emergence of new variants of the virus over time. Infection with the Omicron variant for example has also been shown to cause fever less frequently than the original virus strain.^51, 52^

Our analysis revealed many pathologically high blood pressure values, suggesting a high prevalence of untreated non-communicable diseases within the population. This may reflect difficulties in accessing care and receiving treatment for chronic diseases and may have had implications on adverse outcomes.

We compare complications and outcomes of our cohort to those of cohorts in similar settings. Our choice for comparison includes studies from Yemen, Libya, Sudan, and Somalia, other conflict affected LMICs with similarly fragile health systems.

Of our 1113 included hospitalized patients, a total of 78 (7%) experienced complications, with respiratory complications such as ARDS (3.3%) and respiratory failure (1.8%) being the most common. This number is comparatively low when regarding similar cohorts.^43, 53^ As an example, one retrospective study including 811 hospitalized patients in Libya reported that respiratory distress syndrome occurred in 14.3% of recorded patients.^43^ Such differences in complication rates and adverse outcomes between settings are likely multifactorial, however vaccination status, access to confirmatory testing, oxygen therapy for hypoxaemic patients, prompt administration of steroids for patients requiring oxygen, close nursing monitoring, prone positioning physiotherapy techniques, adequate management of co-morbidities and coexisting pathologies, are all clinical aspects which are likely to vary significantly between settings and health care facilities and will have an impact on patient disease course.

Our outcome description relies on adverse outcome defined as transfer to a level 3 ICU or death. Though little to no data is available on the further development of transferred patients, anecdotal evidence obtained from discussions with the physicians in charge states that a large proportion of patients died after transfer, so that our outcome approximates the mortality rate of the CTC. We do not have enough information to be able to attribute this high mortality among transferred patients to either their extreme severity, the resources available at the transfer destination or to other factors.

As a result, and because of variations in bed capacity and admission criteria, comparison of our outcomes with mortality rates from other studies should be interpreted with care. A total of 165 (14.8%) patients experienced an adverse outcome, which is low compared to similar cohorts. While the Libyan study found a mortality rate of 12.3%, other studies for example in Yemen, Somalia and Sudan described mortality rates of 35, 22 and 21% respectively.^43, 48, 53, 54^ Another survival analysis performed on 131 patients hospitalized in the main hospital in Mogadishu even documented a mortality rate of 40%.^55^

The proportion of adverse outcomes in our study population is low even in comparison with studies from high income countries. One large-scale multicenter observational study including almost half a million hospitalized COVID-19 patients from 49 different countries for example, showed a mortality of 20%.^45^ A further multicenter observational study conducted in the United Kingdom with over 20.000 hospitalized COVID-19 patients, documented a mortality rate of 32%.^46^

Regarding predictive factors for an unfavorable outcome, we identified gender, higher age, O2 flow at admission, lymphopenia, anemia and an O2 saturation of under 94% as variables associated with adverse outcome. This is in line with the current literature.^18-22, 24, 56, 57^ In our univariate analysis we also identified admission status and elevated WBC as factors associated with an adverse outcome. These were not included in the multivariate analysis due to strong correlation with other variables.

The proportion of vaccinated individuals in our cohort was low, with a total of 52 vaccinated patients, corresponding to 17% of patients admitted after vaccination started. This is in line with the current vaccination coverage in Afghanistan. As of July 2022, approximately 13% of the population were fully vaccinated (two doses when required for a specific type of vaccine).^1^ Previous studies have identified supply shortage, insufficient cold chain infrastructure, geographical barriers, political instability and vaccine hesitancy among the population caused by mistrust toward the government and lack of health education as causes of low vaccination coverage in Afghanistan.^58-61^

Self-reported COVID-19 vaccination status was not found to be significantly associated with a better outcome but given the low sample size the power of analysis for this indicator is expected to be very low and should thus be interpreted with caution.

### Strengths and limitations

The unique context of our study location differentiates us from other settings. For the past 40 years Afghanistan has been in almost permanent conflict of fluctuating intensity. The collapsing economy, displacement of approximately four million civilians and further political turmoil acutely seen since July/August 2021 have deteriorated an already struggling healthcare infrastructure, which suffers from a lack of emergency care services, equipment, medication and personnel.^58^ There are currently estimated to be only 1.9 physicians per 10 000 people in the country.^35^ The current pandemic was anticipated to further deteriorate the health care system.^35^

Despite the urgent need for evidence on the evolution of the pandemic to help evaluate and prioritize the most pressing challenges, reliable data is sparse and national data that is recorded is difficult to analyze given lack of completeness, lacking geographical coverage and lack of contextual information limiting interpretability. The national COVID-19 surveillance system suffers from wide-spread under-reporting and a lack of resources for laboratory confirmation and sequencing. In addition, many COVID-19 related deaths are thought to have occurred in the community, not captured by mortality surveillance.^34^ A survey performed in 2020 didn’t provide any insights into COVID-19 related mortality.^62^ In this difficult context, our study provides insights into the in-hospital rate of adverse outcomes and the risk factors associated with severe COVID-19 in MSF’s patient cohort in Herat. It is, however, to be noted that the resources available at an MSF-led facility are more important, specifically in terms of human resources as well as supply of essential drugs and biomedical equipment including oxygen therapy devices, which we infer has likely decreased the rate of adverse outcomes when compared to local hospitals lacking funds, resources, and international support.

Another strength of our analysis lies in the large cohort of over 1000 patients, which ensures adequate statistical power. Furthermore, the use of a database with a web-interface provided capacity for real-time data entry, facilitating access to and real-time monitoring of essential patient indicators and thus allowing not only medico-operational monitoring but also regular remote quality checks by the co-investigators at Epicentre. In addition to regular exchange between the medical personnel at the study site and the investigators this allowed for continuous monitoring and high data reliability.

An important limitation is data completeness. Data collection was performed in a patient treatment setting from clinical files, and in times of high patient load it was common for physicians to skip variables that were not seen to be of immediate clinical relevance. Some of our variables, such as comorbidities, were collected at two time points, at admission and at discharge. Inconsistencies revealed weaknesses during data collection. Thus, several particularly incomplete or inconsistent variables were excluded from our analysis. Although case definitions for severity of disease was generally well understood in this project, the fast evolution of the disease in certain patients made a differentiation between severe and critical states challenging (e.g., for patients who required a gradual increase of O2 flow during the admission).

The proportion of patients who developed adverse outcome was the highest during the third wave, June to August 2021. This overlaps with the time of closure of Shaidayee hospital, the only other CTC in Herat, and equipped with mechanical ventilators. This led to the admission of critically ill patients to the MSF CTC who would otherwise have been referred to Shaidayee. Furthermore, during this time Afghanistan experienced a period of seasonal malnutrition, disruptive political changes and an escalation of conflict, which influenced supply chains.^63^ In addition, it is likely the time when the delta variant of SARS-CoV-2 circulated in Afghanistan. Given these and other similar variations and the fact that during peak times bed capacity was reached, it is likely that not only the effective admission criteria but also the case management capacity and available clinical resources per patient varied through time, meaning that 1) our results may not be representative of complication and mortality rates that would have occurred in a setting not subject to these stressors and 2) complication and adverse outcome rates may not be comparable over time.

Though sequencing of the virus was only very rarely performed in Afghanistan, and not available within this project, SARS-CoV-2 variants of concern also spread through Afghanistan and likely lead to an evolution in severity patterns and immune escape. Extensive literature search identified one study focused on sequencing of SARS-CoV-2 in Afghanistan. In this study, the analysis of 122 COVID samples from foreign soldiers which were collected between February and May 2021 resulted in the detection of 20 virus strains belonging to the delta variant.^64^ In June, news articles quoting the ministry of health, spoke of delta causing up to 60% of cases.^65, 66^ Given the proximity and important population fluxes with Iran and the peak of Delta cases that is documented there in June 2021^67, 68^, one may conclude that the peak in case severity observed at the CTC during the 3^rd^ wave was also related to this variant. Since the proportion of cases of each wave caused by the different variants is not known it is however not possible to conclude on the quantitative impact the variants had on severity.

As an MSF led hospital the CTC benefited from more resources and consequently better staffing, incorporating extended multi-disciplinary teams providing a coordinated holistic patient care approach including, but not limited to, intensive/critical care doctor and nurse supervision & mentoring of local staff, systematic physiotherapy for all inpatients, psychosocial supports and adequate nutrition in addition to advanced biomedical equipment, quality drugs and training, in contrast to what would usually be expected in a similar context. Concurrently, according to the patient flows established in Herat during most of the study period, many critical patients were admitted to Shaidayee hospital and not treated at the MSF CTC. These two factors mean that the rate of adverse outcome and rates of complications measured in our study are not necessarily representative of the context.

A further limitation of our analysis is that laboratory confirmation was not possible for all patients due to limited capacity. Many patients were thus diagnosed based on clinical criteria and epidemiological context, and sometimes RDTs. This was the case in particular in earlier waves where access to testing was even scarcer. A separate analysis of all serologically confirmed cases was not conducted due to risk of bias, given that more severe cases were more likely to be lab tested.

### Implications

This study was initiated as a tool to aid the management of COVID-19 patients by MSF staff in different sites by giving real-time access to detailed clinical patient data during a time when treatment guidelines for COVID-19 patients evolved continuously and protocols applicable in resource limited settings needed to be developed from scratch. Automatized aggregated reports that were shared with COVID-19 referents and the local team helped to highlight weak spots almost in real-time, so that improvements could be continuously implemented.

Furthermore, given that in several countries the COVID-19 pandemic led to MSF implementing a level 2 ICU for the first time, an additional aim of this comprehensive database was to evaluate the infrastructural quality of care and provide lessons learned to guide future emergency response interventions.

Our analysis could contribute to the creation of a risk score for severe disease outcomes to be considered for further outbreaks. The variables included in our analysis are all easily accessible and inexpensive, so adapted to a context with limited resources.

Overall, this study is among the few that longitudinally describe hospitalized COVID-19 patients, their risk factors, complications, and outcomes in a severely conflict affected LMIC setting. Despite the inherent limitations arising from scarce resources, a complex and quickly changing environment, and challenges related to the representativeness of our cohort of patients, our results show that indicators associated with adverse outcome of COVID-19 in Afghanistan are similar to those found in other settings.

## Data Availability

Due to the nature of the study and the potential ease of identification of research participants, publication of the data underlying this study is subject to legal and ethical restrictions. The minimal data set underlying the findings of this study are available on request, in accordance with the legal framework set forth by Médecins Sans Frontières (MSF) data sharing policy (Karunakara U, PLoS Med 2013). MSF is committed to sharing and disseminating health data from its programs and research in an open, timely, and transparent manner in order to promote health benefits for populations while respecting ethical and legal obligations towards patients, research participants, and their communities. The MSF data sharing policy ensures that data will be available upon request to interested researchers while addressing all security, legal, and ethical concerns. All readers may contact the generic address or the corresponding authors to request the data.

## List of abbreviations

ARDS: Acute respiratory distress syndrome
BP: Blood pressure
CI: Confidence interval
COVID (−19): Coronavirus disease
CRP: C-reactive protein
CTC: COVID-19 treatment center
HIV: Human immunodeficiency virus
HR: Hazard ratio
ICU: Intensive care unit
IQR: Interquartile range
LMIC: Low- to middle-income country
MSF: Médecins Sans Frontières
PCR: Polymerase chain reaction
REDCap: Research Electronic Data Capture
WBC: White blood cell
WHO: World Health Organization

## Author contributions

MB, EP and FF designed the study. FF implemented and oversaw the data collection and performed regular quality control and follow-up. AK, MB, EP and FF performed the analysis. HH, SS, BM, GM, AA, AHT, BB, NP and MAZK provided. AK and FF wrote the first draft. All authors contributed to the final version.

## Acknowledgements

This study was funded by MSF. The study protocol is based on a generic protocol developed by Helena Huerga, Sarala Nicholas, Mathieu Bastard, Maria Lightowler, Jeanne Haidar and Elisabeth Poulet at Epicentre. The authors would like to thank all involved health care and data administration staff in Afghanistan for their great work and dedication.

## Supplement

**Supplementary Table 1:** Data collected at admission, discharge and during the patient’s stay.

|  | Admission | Discharge | During stay |
| --- | --- | --- | --- |
| **Patient demographics** | X | - | - |
| **Comorbidities** | X | X | - |
| **Symptoms** | X | X | - |
| **Clinical examination** | X | - | - |
| **Recent medication** | X | - | - |
| **COVID-19 vaccination status** | X | - | - |
| **Oxygen therapy** | X | X | X<sup>1</sup> |
| **Supportive medication** | X | X | - |
| **Laboratory analyses** | (X)* | (X)* | (X)* |
| **Vital signs** | X | (X)* | X<sup>2</sup> |
| **COVID-19 testing** | (X)* | - | (X)* |
| **Supplementary diagnostics** | (X)* | - | - |
| **Complications** | - | X | - |
| **Outcome** | - | X | - |

<sup>1</sup>As part of vital sign collection
<sup>2</sup>vital signs are collected multiple times daily, but only two values per day are entered into the database
\* Upon physicians' discretion and availability

## References

1. Organization WH. WHO Coronavirus (COVID-19) Dashboard, https://covid19.who.int/region/emro/country/af x(accessed 18.07.2022 2022).

2. Wu Z and McGoogan JM. Characteristics of and Important Lessons From the Coronavirus Disease 2019 (COVID-19) Outbreak in China: Summary of a Report of 72314 Cases From the Chinese Center for Disease Control and Prevention. JAMA 2020; 323: 1239–1242. 2020/02/25. DOI: 10.1001/jama.2020.2648.

3. Organization WH. Living guidance for clinical management of COVID-19, https://apps.who.int/iris/bitstream/handle/10665/349321/WHO-2019-nCoV-clinical-2021.2-eng.pdf (2021, accessed 10.08.2022 2022).

4. Barks P and Finger F. Covidestim. Epicentre, GitHub, 2020.

5. da Rosa Mesquita R, Francelino Silva Junior LC, Santos Santana FM, et al. Clinical manifestations of COVID-19 in the general population: systematic review. Wien Klin Wochenschr 2021; 133: 377–382. 2020/11/27. DOI: 10.1007/s00508-020-01760-4.

6. Alimohamadi Y, Sepandi M, Taghdir M, et al. Determine the most common clinical symptoms in COVID-19 patients: a systematic review and meta-analysis. J Prev Med Hyg 2020; 61: E304–E312. 2020/11/06. DOI: 10.15167/2421-4248/jpmh2020.61.3.1530.

7. Yang X, Yu Y, Xu J, et al. Clinical course and outcomes of critically ill patients with SARS-CoV-2 pneumonia in Wuhan, China: a single-centered, retrospective, observational study. Lancet Respir Med 2020; 8: 475–481. 2020/02/28. DOI: 10.1016/S2213-2600(20)30079-5.

8. Tang N, Li D, Wang X, et al. Abnormal coagulation parameters are associated with poor prognosis in patients with novel coronavirus pneumonia. J Thromb Haemost 2020; 18: 844–847. 2020/02/20. DOI: 10.1111/jth.14768.

9. Pan L, Mu M, Yang P, et al. Clinical Characteristics of COVID-19 Patients With Digestive Symptoms in Hubei, China: A Descriptive, Cross-Sectional, Multicenter Study. Am J Gastroenterol 2020; 115: 766–773. 2020/04/15. DOI: 10.14309/ajg.0000000000000620.

10. AbuRuz S, Al-Azayzih A, ZainAlAbdin S, et al. Clinical characteristics and risk factors for mortality among COVID-19 hospitalized patients in UAE: Does ethnic origin have an impact. PLoS One 2022; 17: e0264547. 2022/03/03. DOI: 10.1371/journal.pone.0264547.

11. Pya Y, Bekbossynova M, Gaipov A, et al. Mortality predictors of hospitalized patients with COVID-19: Retrospective cohort study from Nur-Sultan, Kazakhstan. PLoS One 2021; 16: e0261272. 2021/12/23. DOI: 10.1371/journal.pone.0261272.

12. Garcia-Vidal C, Cozar-Llisto A, Meira F, et al. Trends in mortality of hospitalised COVID-19 patients: A single centre observational cohort study from Spain. Lancet Reg Health Eur 2021; 3: 100041. 2021/04/20. DOI: 10.1016/j.lanepe.2021.100041.

13. Myers LC, Parodi SM, Escobar GJ, et al. Characteristics of Hospitalized Adults With COVID-19 in an Integrated Health Care System in California. JAMA 2020; 323: 2195–2198. 2020/04/25. DOI: 10.1001/jama.2020.7202.

14. Salluh JIF, Burghi G and Haniffa R. Intensive care for COVID-19 in low- and middle-income countries: research opportunities and challenges. Intensive Care Med 2021; 47: 226-229. 2020/11/14. DOI: 10.1007/s00134-020-06285-y.

15. Zhou F, Yu T, Du R, et al. Clinical course and risk factors for mortality of adult inpatients with COVID-19 in Wuhan, China: a retrospective cohort study. Lancet 2020; 395: 1054–1062. 2020/03/15. DOI: 10.1016/S0140-6736(20)30566-3.

16. Macedo A, Goncalves N and Febra C. COVID-19 fatality rates in hospitalized patients: systematic review and meta-analysis. Ann Epidemiol 2021; 57: 14–21. 2021/03/05. DOI: 10.1016/j.annepidem.2021.02.012.

17. Dessie ZG and Zewotir T. Mortality-related risk factors of COVID-19: a systematic review and meta-analysis of 42 studies and 423,117 patients. BMC Infect Dis 2021; 21: 855. 2021/08/23. DOI: 10.1186/s12879-021-06536-3.

18. Petrilli CM, Jones SA, Yang J, et al. Factors associated with hospital admission and critical illness among 5279 people with coronavirus disease 2019 in New York City: prospective cohort study. BMJ 2020; 369: m1966. 2020/05/24. DOI: 10.1136/bmj.m1966.

19. Xie J, Covassin N, Fan Z, et al. Association Between Hypoxemia and Mortality in Patients With COVID-19. Mayo Clin Proc 2020; 95: 1138–1147. 2020/05/08. DOI: 10.1016/j.mayocp.2020.04.006.

20. Chatterjee NA, Jensen PN, Harris AW, et al. Admission respiratory status predicts mortality in COVID-19. Influenza Other Respir Viruses 2021; 15: 569-572. 2021/05/25. DOI: 10.1111/irv.12869.

21. Mejia F, Medina C, Cornejo E, et al. Oxygen saturation as a predictor of mortality in hospitalized adult patients with COVID-19 in a public hospital in Lima, Peru. PLoS One 2020; 15: e0244171. 2020/12/29. DOI: 10.1371/journal.pone.0244171.

22. Altschul DJ, Unda SR, Benton J, et al. A novel severity score to predict inpatient mortality in COVID-19 patients. Sci Rep 2020; 10: 16726. 2020/10/09. DOI: 10.1038/s41598-020-73962-9.

23. Marcos M, Belhassen-Garcia M, Sanchez-Puente A, et al. Development of a severity of disease score and classification model by machine learning for hospitalized COVID-19 patients. PLoS One 2021; 16: e0240200. 2021/04/22. DOI: 10.1371/journal.pone.0240200.

24. Anai M, Akaike K, Iwagoe H, et al. Decrease in hemoglobin level predicts increased risk for severe respiratory failure in COVID-19 patients with pneumonia. Respir Investig 2021; 59: 187–193. 2020/12/08. DOI: 10.1016/j.resinv.2020.10.009.

25. Zhang J, Wang X, Jia X, et al. Risk factors for disease severity, unimprovement, and mortality in COVID-19 patients in Wuhan, China. Clin Microbiol Infect 2020; 26: 767–772. 2020/04/19. DOI: 10.1016/j.cmi.2020.04.012.

26. Pyramid P. Population Pyramids of the World from 1950 to 2100, https://www.populationpyramid.net/world/2019/ (2019, accessed 10.08.2022 2022).

27. Ritchie H and Roser M. Age structure, https://ourworldindata.org/age-structure (2019, accessed 10.08.2022 2022).

28. Favas C, Jarrett P, Ratnayake R, et al. Country differences in transmissibility, age distribution and case-fatality of SARS-CoV-2: a global ecological analysis. Int J Infect Dis 2022; 114: 210–218. 2021/11/09. DOI: 10.1016/j.ijid.2021.11.004.

29. Federation ID. IDF Diabetes Atlas, https://diabetesatlas.org/atlas/tenth-edition/ (2021, accessed 10.08.2022 2022).

30. Organization WH. Noncommunicable diseases, https://www.who.int/news-room/fact-sheets/detail/noncommunicable-diseases (2021, accessed 10.08.2022 2022).

31. Thienemann F, Ntusi NAB, Battegay E, et al. Multimorbidity and cardiovascular disease: a perspective on low- and middle-income countries. Cardiovasc Diagn Ther 2020; 10: 376–385. 2020/05/19. DOI: 10.21037/cdt.2019.09.09.

32. Prabhakaran D, Anand S, Watkins DA, et al. Cardiovascular, Respiratory, and Related Disorders: Key Messages and Essential Interventions to Address Their Burden in Low- and Middle-Income Countries. In: rd, Prabhakaran D, Anand S, et al. (eds) Cardiovascular, Respiratory, and Related Disorders. Washington (DC), 2017.

33. Mousavi SH, Shah J, Giang HTN, et al. The first COVID-19 case in Afghanistan acquired from Iran. Lancet Infect Dis 2020; 20: 657–658. 2020/03/28. DOI: 10.1016/S1473-3099(20)30231-0.

34. Nemat A and Asady A. The Third Wave of the COVID-19 in Afghanistan: An Update on Challenges and Recommendations. J Multidiscip Healthc 2021; 14: 2043–2045. 2021/08/12. DOI: 10.2147/JMDH.S325696.

35. Shah J, Karimzadeh S, Al-Ahdal TMA, et al. COVID-19: the current situation in Afghanistan. Lancet Glob Health 2020; 8: e771–e772. 2020/04/06. DOI: 10.1016/S2214-109X(20)30124-8.

36. Cousins S. Afghanistan braced for second wave of COVID-19. Lancet 2020; 396: 1716–1717. 2020/11/30. DOI: 10.1016/S0140-6736(20)32529-0.

37. Asady A, Sediqi MF and Habibi SS. The Fourth Wave of the COVID-19 in Afghanistan: The Way Forward. Infect Drug Resist 2022; 15: 3369–3371. 2022/07/06. DOI: 10.2147/IDR.S365868.

38. Authority NSaI. Estimated Population of Afghanistan 2021-22, https://web.archive.org/web/20210624204559/https:/www.nsia.gov.af:8080/wp-content/uploads/2021/06/Estimated-Population-of-Afghanistan1-1400.pdf (2021).

39. Harris PA, Taylor R, Minor BL, et al. The REDCap consortium: Building an international community of software platform partners. J Biomed Inform 2019; 95: 103208. 2019/05/13. DOI: 10.1016/j.jbi.2019.103208.

40. Harris PA, Taylor R, Thielke R, et al. Research electronic data capture (REDCap)--a metadata-driven methodology and workflow process for providing translational research informatics support. J Biomed Inform 2009; 42: 377–381. 2008/10/22. DOI: 10.1016/j.jbi.2008.08.010.

41. Rosenthal N, Cao Z, Gundrum J, et al. Risk Factors Associated With In-Hospital Mortality in a US National Sample of Patients With COVID-19. JAMA Netw Open 2020; 3: e2029058. 2020/12/11. DOI: 10.1001/jamanetworkopen.2020.29058.

42. Wang Z, Ye D, Wang M, et al. Clinical Features of COVID-19 Patients with Different Outcomes in Wuhan: A Retrospective Observational Study. Biomed Res Int 2020; 2020: 2138387. 2020/10/09. DOI: 10.1155/2020/2138387.

43. Elhadi M, Momen AA, Alsoufi A, et al. Epidemiological and clinical presentations of hospitalized COVID-19 patients in Libya: An initial report from Africa. Travel Med Infect Dis 2021; 42: 102064. 2021/04/21. DOI: 10.1016/j.tmaid.2021.102064.

44. Di Fusco M, Shea KM, Lin J, et al. Health outcomes and economic burden of hospitalized COVID-19 patients in the United States. J Med Econ 2021; 24: 308–317. 2021/02/09. DOI: 10.1080/13696998.2021.1886109.

45. Kartsonaki C. Characteristics and outcomes of an international cohort of 400,000 hospitalised patients with Covid-19. medRxiv 2021: 2021.2009.2011.21263419. DOI: 10.1101/2021.09.11.21263419.

46. Docherty AB, Harrison EM, Green CA, et al. Features of 20 133 UK patients in hospital with covid-19 using the ISARIC WHO Clinical Characterisation Protocol: prospective observational cohort study. BMJ 2020; 369: m1985. DOI: 10.1136/bmj.m1985.

47. Zhu J, Ji P, Pang J, et al. Clinical characteristics of 3062 COVID-19 patients: A meta-analysis. J Med Virol 2020; 92: 1902–1914. 2020/04/16. DOI: 10.1002/jmv.25884.

48. Hasabo EA, Ayyad FA, Alam Eldeen SAM, et al. Clinical manifestations, complications, and outcomes of patients with COVID-19 in Sudan: a multicenter observational study. Trop Med Health 2021; 49: 91. 2021/11/16. DOI: 10.1186/s41182-021-00382-4.

49. Ben-Tov A, Lotan R, Gazit S, et al. Dynamics in COVID-19 symptoms during different waves of the pandemic among children infected with SARS-CoV-2 in the ambulatory setting. Eur J Pediatr 2022 2022/07/02. DOI: 10.1007/s00431-022-04531-7.

50. Ramos-Rincon JM, Cobos-Palacios L, Lopez-Sampalo A, et al. Differences in clinical features and mortality in very old unvaccinated patients (>/= 80 years) hospitalized with COVID-19 during the first and successive waves from the multicenter SEMI-COVID-19 Registry (Spain). BMC Geriatr 2022; 22: 546. 2022/07/01. DOI: 10.1186/s12877-022-03191-4.

51. Petersen MS, S IK, Eliasen EH, et al. Clinical characteristics of the Omicron variant - results from a Nationwide Symptoms Survey in the Faroe Islands. Int J Infect Dis 2022; 122: 636–643. 2022/07/11. DOI: 10.1016/j.ijid.2022.07.005.

52. Vihta KD, Pouwels KB, Peto TE, et al. Omicron-associated changes in SARS-CoV-2 symptoms in the United Kingdom. Clin Infect Dis 2022 2022/08/03. DOI: 10.1093/cid/ciac613.

53. Ahmed MAM, Hussein AM, Abdullahi AAM, et al. Cardiovascular risk factors and clinical outcomes of patients hospitalized with COVID-19 pneumonia in Somalia. Ther Adv Infect Dis 2022; 9: 20499361221095731. 2022/05/03. DOI: 10.1177/20499361221095731.

54. Al-Waleedi AA, Naiene JD, Thabet AAK, et al. The first 2 months of the SARS-CoV-2 epidemic in Yemen: Analysis of the surveillance data. PLoS One 2020; 15: e0241260. 2020/10/30. DOI: 10.1371/journal.pone.0241260.

55. Ali MM, Malik MR, Ahmed AY, et al. Survival analysis of all critically ill patients with COVID-19 admitted to the main hospital in Mogadishu, Somalia, 30 March-12 June 2020: which interventions are proving effective in fragile states? Int J Infect Dis 2022; 114: 202–209. 2021/11/16. DOI: 10.1016/j.ijid.2021.11.018.

56. Niu J, Sareli C, Mayer D, et al. Lymphopenia as a Predictor for Adverse Clinical Outcomes in Hospitalized Patients with COVID-19: A Single Center Retrospective Study of 4485 Cases. J Clin Med 2022; 11 2022/02/16. DOI: 10.3390/jcm11030700.

57. Tan L, Wang Q, Zhang D, et al. Lymphopenia predicts disease severity of COVID-19: a descriptive and predictive study. Signal Transduct Target Ther 2020; 5: 33. 2020/04/17. DOI: 10.1038/s41392-020-0148-4.

58. Essar MY, Hasan MM, Islam Z, et al. COVID-19 and multiple crises in Afghanistan: an urgent battle. Confl Health 2021; 15: 70. 2021/09/19. DOI: 10.1186/s13031-021-00406-0.

59. Essar MY, Wara UU, Mohan A, et al. Challenges of COVID-19 vaccination in Afghanistan: A rising concern. Ethics Med Public Health 2021; 19: 100703. 2021/07/08. DOI: 10.1016/j.jemep.2021.100703.

60. Shoib S, Saleem SM, Essar MY, et al. Challenges faced by healthcare workers in Afghanistan amidst the COVID-19 pandemic and political instability: A call for action. Clin Epidemiol Glob Health 2022; 15: 101050. 2022/05/03. DOI: 10.1016/j.cegh.2022.101050.

61. Nemat A, Bahez A, Salih M, et al. Public Willingness and Hesitancy to Take the COVID-19 Vaccine in Afghanistan. Am J Trop Med Hyg 2021; 105: 713–717. 2021/07/09. DOI: 10.4269/ajtmh.21-0231.

62. Saeedzai SA, Sahak MN, Arifi F, et al. COVID-19 morbidity in Afghanistan: a nationwide, population-based seroepidemiological study. BMJ Open 2022; 12: e060739. DOI: 10.1136/bmjopen-2021-060739.

63. Taconet A. Caring for patients with covid-19 in Afghanistan-another hard winter. BMJ 2022; 376: o175. 2022/01/23. DOI: 10.1136/bmj.o175.

64. Maljkovic Berry I, Hang J, Fung C, et al. Genomic surveillance of SARS-CoV-2 in US military compounds in Afghanistan reveals multiple introductions and outbreaks of Alpha and Delta variants. BMC Genomics 2022; 23: 513. 2022/07/16. DOI: 10.1186/s12864-022-08757-5.

65. Delta Variant Makes 50% of COVID-19 Infections in Afghanistan. TOLOnews, 27.06.2021 2021.

66. News A. COVID-19 Delta variant of COVID-19 spreading rapidly through Afghanistan. ATN News, 26.09.2021 2021.

67. Sheikhi F, Yousefian N, Tehranipoor P, et al. Estimation of the basic reproduction number of Alpha and Delta variants of COVID-19 pandemic in Iran. PLoS One 2022; 17: e0265489. 2022/05/18. DOI: 10.1371/journal.pone.0265489.

68. Yavarian J, Nejati A, Salimi V, et al. Whole genome sequencing of SARS-CoV2 strains circulating in Iran during five waves of pandemic. PLoS One 2022; 17: e0267847. 2022/05/03. DOI: 10.1371/journal.pone.0267847.

